## Supplemental Text for "Early prediction of clinical response to checkpoint inhibitor therapy in human solid tumors through mathematical modeling"

### Supplementary Figures

**Figure S1**.

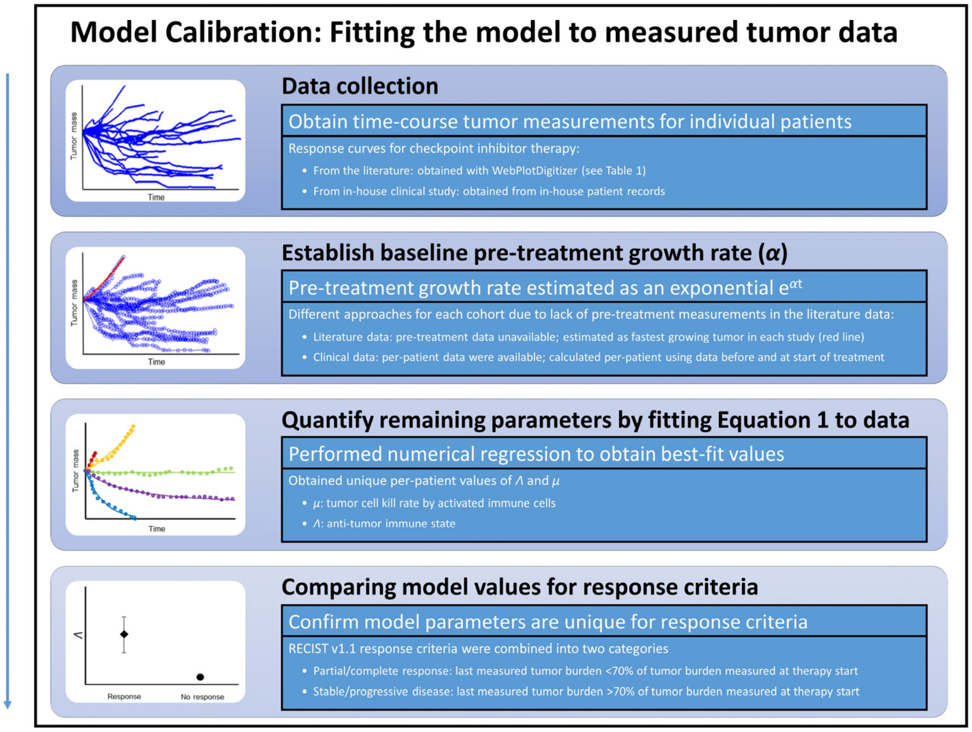


**Figure S1.** Steps for calibration of the mathematical model with clinical data. First, checkpoint inhibitor response curves were extracted from the literature. First, in all cases, immunotherapy treatment began at time $t$ = 0. Second, a tumor specific proliferation constant ($\alpha$) was determined for each cancer type by fitting exponential function ($e^{\alpha t}$) to fastest progressing patient in each clinical trial (red line). Third, individual patient response data were fit to Equation (1) by using the respective α to determine $\Lambda$ and $\mu$. $\Lambda$ and $\mu$ values were then with compared in patients with partial/complete response versus patients with stable/progressive disease after immunotherapy by using the RECIST v1.1 criteria.

**Figure S2**.

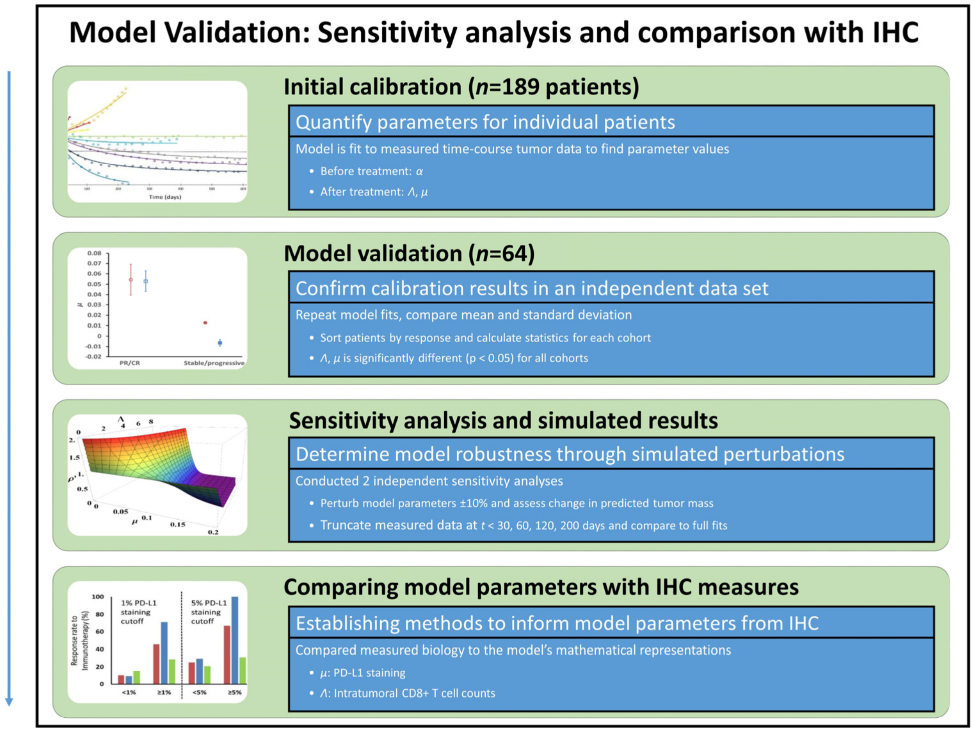


**Figure S2**. Model validation, sensitivity studies, and comparison of model parameters to IHC measures. Model parameters were obtained from a second in-house patient cohort of patients with NSCLC ($n$ = 64), which were compared to values obtained in the calibration cohort in a validation study. To study the sensitivity of the model to changes in model parameter values, key parameters were perturbed ±10% and the resultant simulated expected tumor burden was compared to measured values pre-perturbation. Tumor burden measures were also truncated, and results of truncated and full dataset model fits were compared. Lastly, the full parameter space of the model was examined. In order to compare model parameters to the underlying biology, model parameters were converted to intratumoral CD8+ lymphocyte counts (for $\Lambda$) and PD-L1 staining (for $\mu$), which were compared to IHC measures obtained from the literature.

### Supplementary Table S1

**Table S1**. Studies used for derivation of pathologic markers of immunotherapy response

| **Reference** (see main text) | **Tumor Type** | **Checkpoint Inhibitor** | **Pathologic Biomarker** | **PD-L1 Staining Cutoff** |
|---|---|---|---|---|
| (13) | Melanoma | Pembrolizumab | CD8+ TILs | N/A |
| (45) | Melanoma | Pembrolizumab | PD-L1 | 1% |
| (39) | UCC | Atezolizumab | PD-L1 | 1%, 5%, 10% |
| (44) | NSCLC, RCC, melanoma, HNSCC, CRC, gastric and pancreatic cancer | Atezolizumab | PD-L1 | 1%, 5%, 10% |
| (3) | Melanoma | Nivolumab | PD-L1 | 5% |
| (38) | RCC | Nivolumab | PD-L1 | 5% |
| (47) | NSCLC, RCC, melanoma, PC, CRC | Nivolumab | PD-L1 | 5% |
| (46) | NSCLC | Atezolizumab | PD-L1 | 1%, 5%, 10% |
| (8) | NSCLC | Nivolumab | PD-L1 | 1%, 5%, 10% |
| (1) | NSCLC | Nivolumab | PD-L1 | 1%, 5%, 10% |
| (48) | Melanoma | Nivolumab | PD-L1 | 5% |
| (40) | Melanoma, RCC, NSCLC, CRC, PC | Nivolumab | PD-L1 | 5% |
| (43) | NSCLC | Pembrolizumab | PD-L1 | 1%, 50% |

RCC: Renal cell carcinoma
UCC: Urothelial cell carcinoma
CRC: Colorectal carcinoma
NSCLC: Non-small lung carcinoma
HNSCC: Head and neck squamous cell carcinoma
PC: Prostate carcinoma
TIL: Tumor-infiltrating lymphocytes
